# Application of a general LLM-based classification system to retrieve information about oncological trials

**DOI:** 10.1101/2024.12.03.24318390

**Authors:** Fabio Dennstädt, Paul Windisch, Irina Filchenko, Johannes Zink, Paul Martin Putora, Ahmed Shaheen, Roberto Gaio, Nikola Cihoric, Marie Wosny, Stefanie Aeppli, Max Schmerder, Mohamed Shelan, Janna Hastings

**Author notes:** Contact, Fabio Dennstädt –.

## Abstract

**Purpose:** The automated classification of clinical trials and medical literature is increasingly relevant, particularly in oncology, as the volume of publications and trial reports continues to expand. Large Language Models (LLMs) may provide new opportunities for automated diverse classification tasks. In this study, we developed a general-purpose text classification framework using LLMs and evaluated its performance on oncological trial classification tasks.

**Methods and Materials:** A general text classification framework with adaptable prompt, model and categories for the classification was developed. The framework was tested with four datasets comprising nine binary classification questions related to oncological trials. Evaluation was conducted using a locally hosted version of Mixtral-8x7B-Instruct v0.1 and three cloud-based LLMs: Mixtral-8x7B-Instruct v0.1, Llama3.1-70B-Instruct, and Qwen-2.5-72B.

**Results:** The system consistently produced valid responses with the local Mixtral-8x7B-Instruct model and the Llama3.1-70B-Instruct model. It achieved a response validity rate of 99.70% and 99.88% for the cloud-based Mixtral and Qwen models, respectively. Across all models, the framework achieved an overall accuracy of >94%, precision of >92%, recall of >90%, and an F1-score of >92%. Question-specific accuracy ranged from 86.33% to 99.83% for the local Mixtral model, 85.49% to 99.83% for the cloud-based Mixtral model, 90.50% to 99.83% for the Llama3.1 model, and 77.13% to 99.83% for the Qwen model.

**Conclusions:** The LLM-based classification framework exhibits robust accuracy and adaptability across various oncological trial classification tasks. The findings highlight the potential of automated, LLM-driven trial classification systems, which may become increasingly used in oncology.

## INTRODUCTION

The volume of published biomedical research has grown substantially over recent decades (1). This substantial increase underscores the challenge of efficiently identifying and synthesizing scientific evidence to inform clinical decision-making. In oncology, one of the most rapidly advancing fields of medicine, randomized controlled trials (RCTs) are considered the gold standard for evidence-based decision-making. Accurate classification of trial data is essential, as cancer diagnoses and treatment strategies often rely on a variety of classification systems. These include tumor staging systems like TNM, molecular and genetic profiling, risk stratification systems such as the Gleason Score for prostate cancer, and general health scales like ECOG or Karnofsky Performance Status. Moreover, the diversity of trial settings and endpoints adds complexity. Depending on the research context, clinical trials may evaluate endpoints such as overall survival, progression-free survival, or quality-of-life measures (e.g., ECOG performance scale).

In this context, manual curation of research evidence is increasingly impractical. Using natural language processing (NLP) approaches to automatically classify clinical trials or answer certain questions about them has therefore been of longstanding interest (2) (3). Due to the large number or trials published every year it is virtually impossible to keep track. On ClinicalTrials.gov alone there are roughly half a million registered studies, most of them interventional studies, with oncology accounting for about a fifth (4) (5).

An automated system for classifying oncological trials would be highly valuable. Such a system could substantially support systematic literature reviews and meta-analyses, facilitating the maintenance of up-to-date investigations as part of concepts like living systematic reviews (6) and living guidelines (7).

Current solutions like Trialstreamer (8) using both machine learning (ML) and rule-based approaches are being used for RCTs. Such systems have shown promising results in extracting defined PICO (population, intervention, comparator and outcome) characteristics about RCTs from scientific abstracts.

Using optimization techniques like fine-tuning of ML models and iterative optimization, current solutions can already classify trials with high accuracy into established categories.

A classification system that is not only highly accurate but also adaptable to various classification tasks would be highly valuable. Classifying a trial essentially involves answering a predefined categorical question about the trial (e.g., *“Is this trial in the field of oncology? Yes or No?”*). The concept of a flexible *general classifier* is to provide answers to any question without requiring additional development, such as task-specific optimization or fine-tuning.

Successful development of such a system has been limited with classical NLP techniques for text classification like bag-of-words models or term-frequency-inversed document frequency (TF-IDF) (9). In contrast, traditional ML algorithms (e.g., support vector machines or random forests) often require extensive feature engineering and struggle to generalize across diverse tasks or datasets (10).

Large Language Models (LLMs) are trained on extensive datasets and provide innovative ways to interpret data, access knowledge, and answer complex questions (11). Recent advancements in LLM performance have enabled a range of new applications in medical and clinical knowledge generation and analysis. LLMs have demonstrated substantial potential across various tasks, including medical question-answering (12), summarization of clinical text (13), data extraction from unstructured text (14), and generating high-quality medical writing (15). In one of our studies, we developed a framework for automated LLM-based title-and-abstract screening, showing promising results across various biomedical domains (16). Title-and-abstract screening to check for the relevance of a publication is a specific classification application (*“Is this publication relevant for the topic? Yes or No?”*).

Based on our previous work, we tested the feasibility of developing a *general classifier* to answer various questions about oncological trials based on corresponding text (like the abstract of the publication).

## METHODS

### Development of a general LLM-based classification framework

We developed a straightforward, general approach for using LLMs to classify any given text into user-defined categories. An overview of the concept is shown in **Figure 1**.

**Figure 1:**
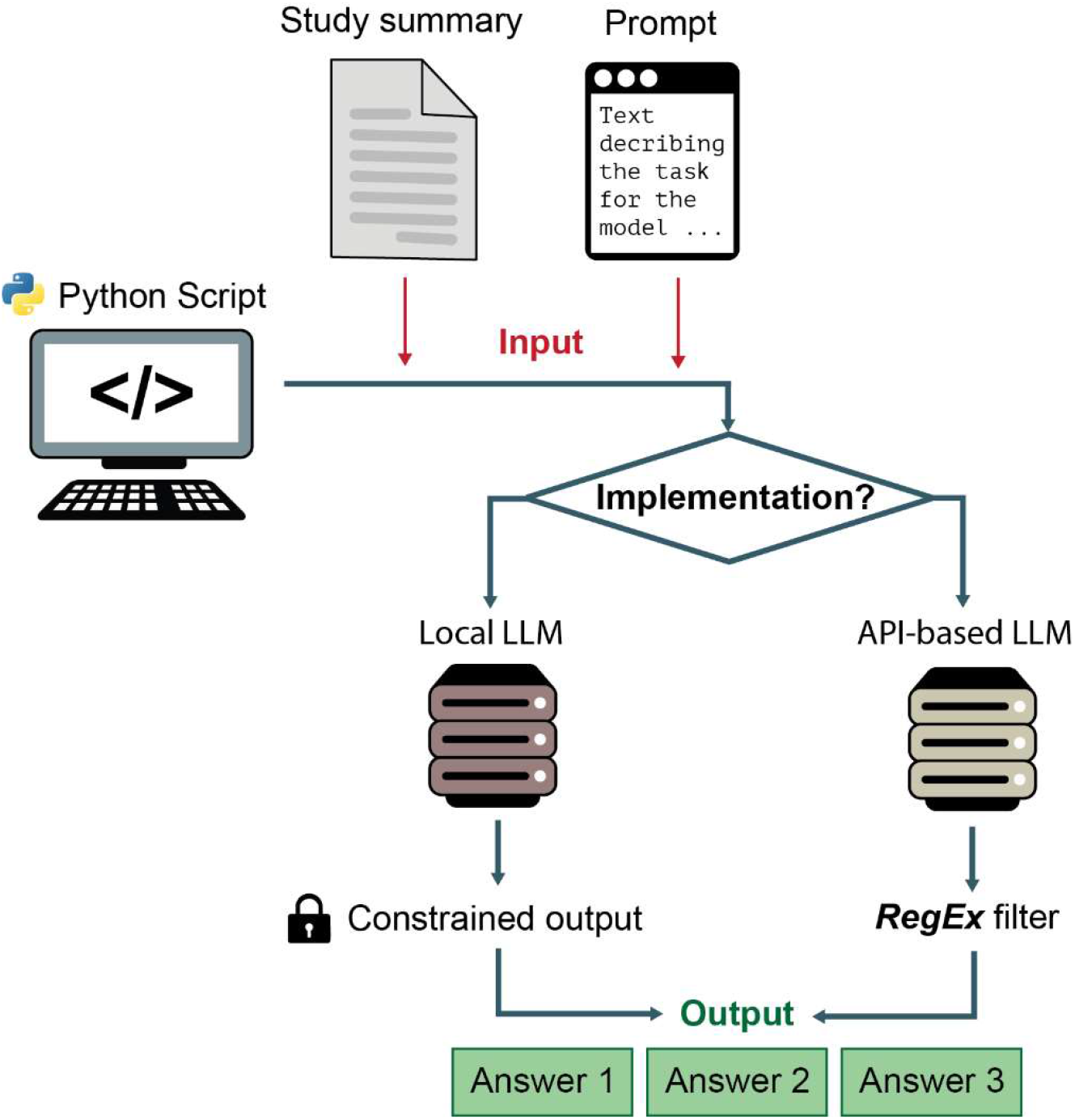
Schematic illustration of the *general classifier* framework. The output of the LLM is either directly constrained (if run on local hardware) or can be analyzed using regular expressions.

The approach follows these steps:

- Defining categories: The categories for classification are specified by the user.
- Text input: The model receives a text string for the classification task, which includes:
- A text description of the classification task and the predefined categories.
- The text to be classified, based on the task or question to be answered.
- LLM: An LLM is run to perform the classification and is run either locally or called via API. The model generates a text output based on the input.
- Category determination: The output is either directly constrained to ensure it matches one of the defined categories, or it is analyzed with methods such as regular expressions to determine which category the model has selected.

Constraining the output of an LLM has the advantage, that the classifier will always give a valid response as it must select one of the defined categories. Constraining can be done using local implementation of libraries like Guidance (17) or Outlines (18). However, as libraries need to be directly implemented when performing output calculations, the model is run on local hardware. This can be a problem if state-of-the-art models are used that require adequate hardware infrastructure and considerable computing resources. To implement and test more resource-intense models that could not be run on our local hardware (i.e., two Nvidia RTX6000 GPUs, each of which with 48GB of RAM), we used the cloud service DeepInfra (19). Since direct constraining of the output is not possible if models are called via API in a cloud-based approach, we used in these cases regular expressions to check for the occurrence of the defined categories in the output of the model. However, in this case the output of an LLM is not principally limited. For the experiments using the cloud computing solution, it is therefore possible that the *general classifier* gives an invalid output if the answer does not contain a text string corresponding to one of the defined categories.

A Python script was created to implement this process, including the direct evaluation of the approach and a basic user interface (UI). The script is publicly available at https://github.com/med-data-tools/general-classifier.

### Large Language Models

Different models were tested for the *general classifier* framework to investigate the approach with various LLMs that are also diverse regarding design and training data. The following three open-source models were used in the experiments:

- Mixtral-8x7B-Instruct-v0.1 is a pretrained generative Mixture of Experts LLM developed by Mistral AI (20), (21). It is a powerful model reported to outperform gpt-3.5-turbo, Claude-2.1, Gemini Pro, and Llama 2 70B-chat on human benchmarks. The script with the model was run both on our local hardware (using a GPTQ version of the model (22)), as well as on cloud computing (23). It should be noted that some small variations in output between local and cloud-based model variant are to be expected due to differences in model versions (GPTQ vs. base), hardware configurations, software environments, and drivers.
- Meta-Llama-3.1-70B-Instruct is a powerful model published in July 2024 by Meta AI (24). The model is specifically tuned for improved reliability and safety in real-world applications. It builds on the Llama 3 architecture with fine-tuning that optimizes response accuracy, reduces biases, and increases usability, targeting better alignment with user intent and enhancing adaptability across various tasks. The model was run using cloud computing (25).
- Qwen2.5-72B-Instruct is Alibaba’s advanced LLM (26) designed for high-performance instruction-following with a focus on accuracy and depth in complex tasks. With 72 billion parameters, this model improves understanding and response generation, aiming for versatility across professional and technical domains. The model was run using cloud computing (27).

## Datasets and questions for the evaluation

For the evaluation, several datasets created by human annotators classifying oncological trials were used. Four different datasets with eleven classification tasks were used in the study. The first two datasets had been annotated as part of previous projects of our group (28) (29) (30) (31).

The third and fourth dataset were created as part of an ongoing systematic review of clinical trials on metastatic castration-resistant prostate cancer (mCRPC) (32). Briefly, the trials in the third and fourth dataset were independently annotated by two researchers (SA and MW) with a third researcher (JH) making a final decision in case of disagreement. All datasets had been created after publication of the LLMs, ensuring that the datasets were not part of the training data of the models.

Dataset 1 contains the abstracts of 899 trials with labels whether they are oncological trials and/or RCTs.

Dataset 2 contains the abstracts of 600 oncological trials with labels whether patients with non-metastatic and/or metastatic disease were included and labels about the type of primary cancer.

Dataset 3 contains abstracts with accompanying information about 144 oncological trials and the label whether it is a trial about patients with mCRPC.

Dataset 4 contains abstracts with accompanying information about 64 oncological trials and the label of whether the trial investigated a therapeutic intervention.

To ensure clear binary categories, the datasets were designed or adapted by breaking down compound classifications or questions into corresponding binary questions answerable with ‘true/false’ or ‘yes/no’. **Figure 2** illustrates how more complex questions in Dataset 1 and Dataset 2 were simplified into corresponding binary questions.

**Figure 2:**
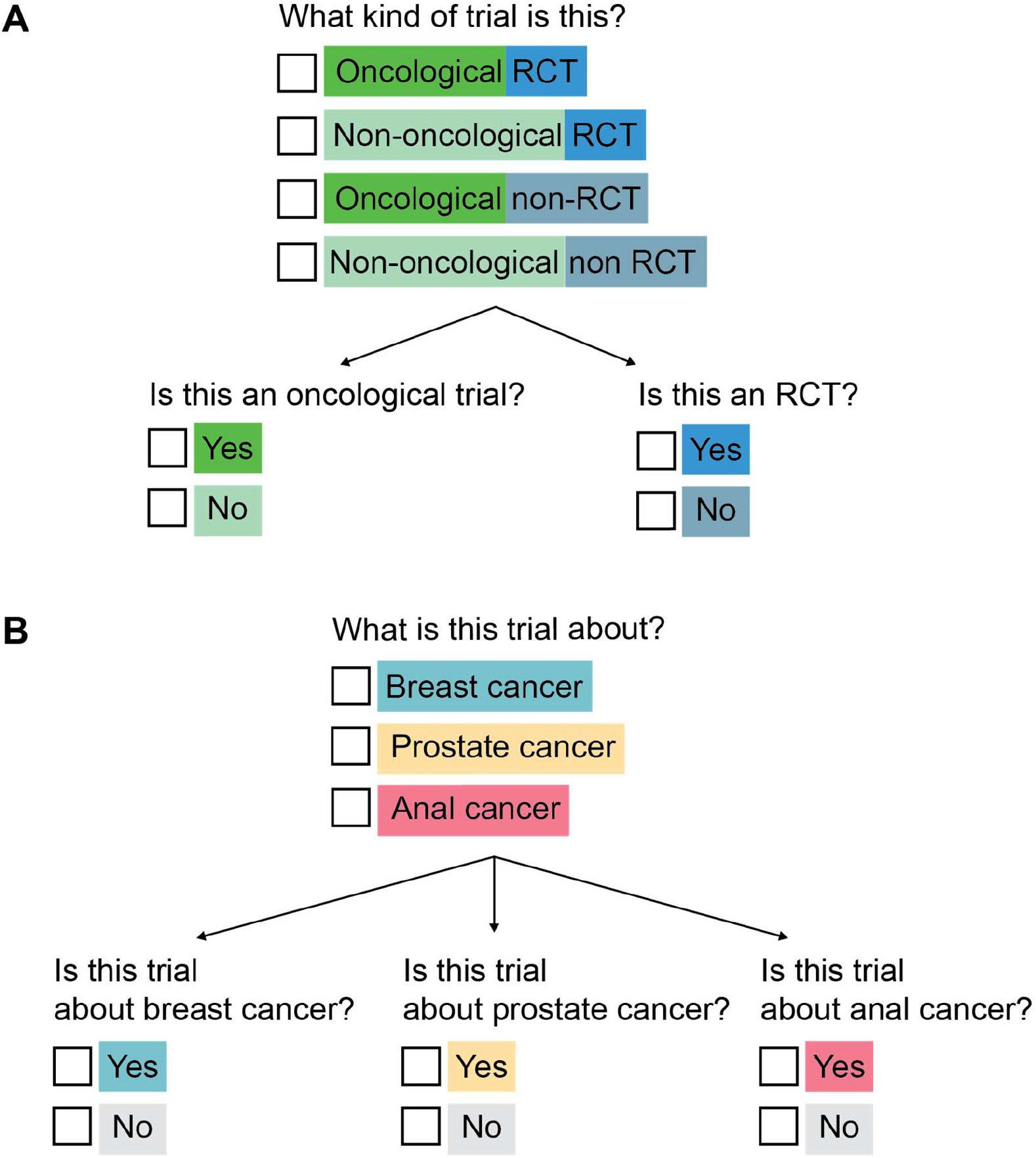
Schematic illustration of breaking down of compound classifications or questions into corresponding binary questions answerable with ‘true/false’ or ‘yes/no’.

Breaking down the questions this way simplifies the task and defines clear concepts with mutually exclusive binary categories. This is important for the classification as only a single category is selected by the *general classifier*. Furthermore, it allows the calculation of classification metrics like accuracy, precision, recall and F1-score.

However, it should be noted that the script in general works also with more than two categories and can also perform several classification tasks in the same run on one dataset.

In total nine binary questions labeled with “true” and “false” were provided in the datasets on which the classifier was tested (**Figure 3**).

**Figure 3:**
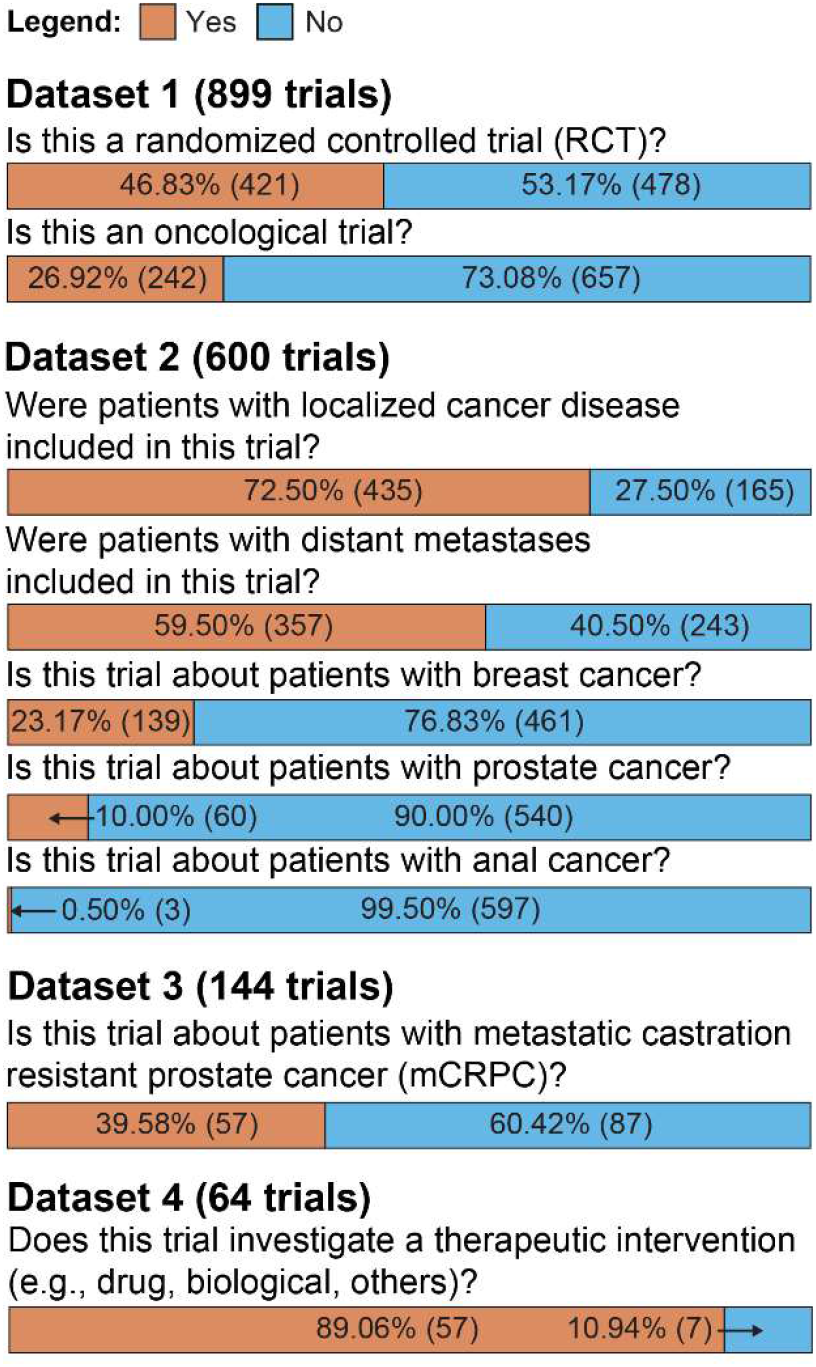
Questions/classification tasks in the four datasets used in the study with the proportion of answers labeled with ‘yes/true’ and ‘no/false’.

The datasets used in the study are provided at *https://github.com/med-data-tools/general-classifier*.

## Prompts for classification tasks

The relevant flexible factor changed by the user of the *general classifier* is the input text / prompt given to the LLM explaining the classification task.

The prompts were created in a non-systematic way following a basic structure containing a brief Instruction (“You are a helpful classifier”), followed by a brief explanation that a question about a clinical trial is to be answered followed by the text for classification and the question. Furthermore, the beginning of the answer to that question was also included, so that a reasonable context is already given before the LLM provides the answer.

As an example, the prompt for the question on RCTs was the following:

“*INSTRUCTION: You are a helpful classifier. You are given the abstract of a scientific publication and you have to decide whether it is of a Randomized Controlled Trial (Answer is ‘TRUE’) or it is not of a Randomized Controlled Trial (Answer is ‘FALSE’). QUESTION: The abstract is ‘[TEXT]’. ANSWER: The correct answer for this abstract is ‘‘*“

(The [TEXT] string is replaced with the abstract of the trial to be classified).

The settings and prompts used in the experiments for all the questions are provided at *https://github.com/med-data-tools/general-classifier*.

As it has been shown, multiple strategies exist to improve outputs from LLMs and enhance performance in LLM-based classification tasks (33). However, the goal of the *general classifier* framework is to create a flexible tool that could be applied to new questions without requiring extensive prompt optimization. Consequently, no systematic or unsystematic optimization of the prompts was conducted for this study.

## Evaluation, statistical analyses

The performance of the classifier, based on the model used, was evaluated using accuracy, precision, recall, and F1-score. In cases where cloud computing was utilized (using non-constrained output), the number of invalid responses produced by the classifier was also measured. These metrics were calculated for individual datasets as well as for the combined data, aggregating all classification cases.

## RESULTS

### Performance of locally run Mixtral-8x7B-Instruct-v0.1-GPTQ (Constrained output)

Since the output of the locally run Mixtral-8x7B-Instruct v0.1-GPTQ was constrained, it always gave valid responses for the classification.

**Figure 4** shows the confusion matrices of the general LLM-based classifier on all nine questions across the four datasets using the locally run model.

**Figure 4:**
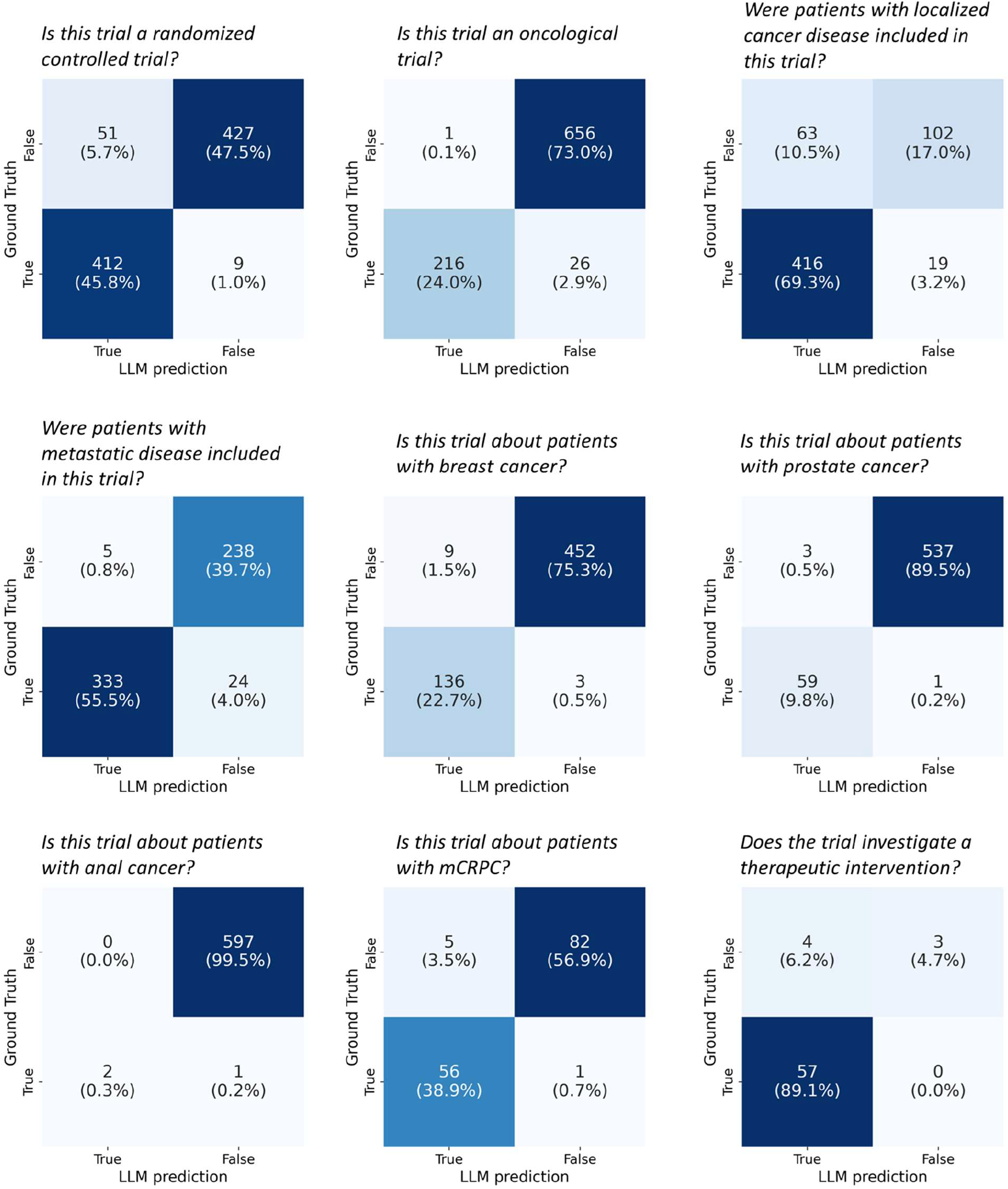
Confusion matrices for the performance of the *general classifier* using the local Mixtral-8x7B-Instruct-v0.1-GPTQ model.

In comparison, using the cloud-run version with output analysis using regular expressions, the Mixtral8x7B model returned some invalid responses (1.56%) on the question *“Were patients with localized cancer disease included in this trial?”*, while valid responses were obtained for all the other questions. The Qwen-2.5.-72B-Instruct model provided some invalid responses (0.33 to 0.11%) on 4 of the 9 questions. The Llama-3.1-Instruct-v0.1 always returned valid responses. An overview is provided in **Supplementary Table 1**.

**Table 1** provides an overview of the overall classification metrics using the different models.

**Table 1.** Classification metrics of different LLMs on all nine questions. Values for best-performing model in bold.

|  | *Local Hardware* | *Cloud computing* |  |  |
| --- | --- | --- | --- | --- |
|  | **Mixtral-8x7B-Instruct-v0.1** | **Mixtral-8x7B-Instruct-v0.1** | **Llama3.1-70B-Instruct** | **Qwen2.5-72B-Instruct** |
| Accuracy | 95.51% | 95.99% | **96.98%** | 94.56% |
| Precision | 92.29% | **94.76%** | 94.51% | 94.26% |
| Recall | 95.26% | 93.90% | **97.12%** | 90.10% |
| F1 | 93.75% | 94.33% | **95.80%** | 92.13% |

The lowest performance across all models was observed for the question “*Were patients with localized cancer disease included in this trial?*” with accuracies of 86.33% (Mixtral local), 85.49% (Mixtral cloud), 90.50% (Llama3.1), and 77.13% (Qwen2.5). For the remaining eight questions, all models achieved accuracies above 90%, with the Llama3.1 model reaching accuracies greater than 95%.

An overview of the classification metrics of the models on the individual questions is provided in **Supplementary Table 2**.

## DISCUSSION

### Comparison with classical NLP approaches

Recent technological advancements led to improved performance of LLMs. Apart from well-known examples like general chat systems (e.g., ChatGPT), successful medical applications were demonstrated for various tasks like summarization, translation, or data extraction (13) (14).

Text classification is a foundational task in NLP with proven success in well-defined use cases. As the volume of medical literature, particularly in oncology, continues to grow, automated systems for the classification and analysis of scientific publications are increasingly important. Our findings demonstrate that a general-purpose classification system using modern LLMs can effectively analyze clinical trials by accurately answering related questions. Unlike traditional ML and NLP approaches, this *general classifier* does not require task-specific adaptations (e.g., fine-tuning or retraining), beyond providing the appropriate text prompt for the classification task.

In a previous study we used a RoBERTa-based transformer model to classify the dataset on RCTs (*Is this a randomized controlled trial (RCT)?*) after training the model using a random 85:15 split. The system achieved an accuracy of 94%, precision of 100%, recall of 92% and an F1-score of 96% (31), which is comparable to the performance of the *general classifier* using the Llama 3.1 model (see also **Supplementary Table 2**).

Also on other datasets tested in some of our previous studies (31) (34) the *general classifier* achieved good performance with results comparable to optimized ML- and rule-based approaches. The results of our study show that an LLM-based *general classifier* can be applied to various questions on oncological trials with high rates of accuracy.

There are numerous potential applications and use cases for such a system, including citation screening, collection of trial eligibility criteria (see also (35)), and grouping of trials based on specific criteria.

Automating these tasks can aid in systematic literature reviews, meta-analyses, knowledge collection, and trial database management. While the approach used in the LLM-based *general classifier* was applied to oncological trials in this study, it could be applied to other domains as well. The system can, in principle, classify any given text into any set of user-defined categories, making it highly adaptable to a wide range of use cases.

While the framework itself is general, depending on the specific use case, optimization techniques such as fine-tuning LLMs, in-context learning, or prompt optimization could still be applied. However, the fact that the system achieved high performance without relying on such techniques suggests its potential for widespread applicability and utility.

Rather than relying on widely used, powerful LLMs provided by private companies, such as GPT-4 by OpenAI or Claude 3.5 by Anthropic, we deliberately chose to use open-source models. While these models are still resource-intensive (which is why we needed to rely on cloud computing services), they can, in principle, be deployed on local hardware in a private environment. Relying on LLMs that are accessible only through APIs, effectively “black boxes” controlled by private companies, limits transparency and could hinder the accessibility of the system. For academic biomedical research, it is preferable for literature or trial classification systems to be independent of proprietary LLMs provided by tech companies, ensuring greater transparency and autonomy.

While the results of our study are promising, there are several limitations. The approach itself is resource-intensive and requires adequate hardware to function effectively. Additionally, we tested the method on a limited set of nine questions, which can be answered by binary ‘True/False’ answers. As a result, the generalizability of the findings may be restricted. Using binary categories, rather than multiple categories, enables clearer definitions and facilitates better interpretation of results. In general, classification tasks with multiple categories can be broken down into binary categories (see Figure 2). Overall, selecting a single correct class from multiple choices is meaningful only when the categories are mutually exclusive and free from semantic overlap.

It is important to note that evaluating an LLM-based classification system requires the use of new, unseen datasets. LLMs which are trained on extensive internet text data (11), should not have previously encountered the datasets used in such evaluations. The presented *general classifier* framework itself, also has limitations. Although it can be used to answer questions about oncological trials, it is currently limited to questions with categorical answers. For example, questions requiring numerical responses, such as *“How many patients were included in this trial?”*, cannot be answered in its present form. Additionally, the framework is confined to text-based data, meaning that information presented in non-textual forms, such as figures and tables, cannot be directly incorporated into the classification process. Future developments may explore the use of vision-language models or integrate optical character recognition (OCR) technologies into such a framework, which could enable the inclusion of such visual data (36).

A fundamental limitation of any LLM-based approach is its reliance on the input prompt, which directly influences the results. While the ability to modify the input prompt is a key feature that allows the classification system to be flexible, it also makes the LLM output, and, consequently, the performance of the classifier, difficult to predict. Thus, it is essential to formulate clear, well-defined questions with unambiguous answers. Precise descriptions and clear definitions are required to ensure high performance of a classification system. This may partly explain the slight decline in the performance on the less clearly defined question, “*Were patients with localized disease included in this trial?*”. However, ambiguous definitions would likely also affect human performance, leading to inconsistencies in answering such questions.

Despite these limitations, we believe LLM-based classification systems will play a crucial role in the analysis of medical literature and clinical trials in the future. Oncology might benefit substantially from these advancements, as it involves complex criteria, large datasets, and the nuanced interpretation of clinical evidence to guide decision-making.

## CONCLUSIONS

The *general classifier* developed in our study demonstrated promising results in answering a variety of questions related to oncological trials. It offers a flexible framework for the automatic classification of trials, literature, and other text forms into predefined categories. While there are still some limitations that need to be addressed in future work, we are confident that LLM-based classification systems will become increasingly valuable, particularly in oncological research.

## Data Availability

The source code for the general classifier as well as the datasets used in the experiments are available at https://github.com/med-data-tools/general-classifier

## List of abbreviations

AI: artificial intelligence
API: application programming interface
ISROI: International Society for Radiation Oncology
LLM: Large Language Model
mCRPC: metastatic castration resistant prostate cancer
ML: machine learning
NLP: natural language processing
OCR: optical character recognition
PICO: population, intervention, comparator and outcome
RCT: randomized controlled trial
TF-IDF: term-frequency-inversed document frequency
UI: – user interface

## Code and data availability statement

The source code for the *general classifier* as well as the datasets used in the experiments are available at *https://github.com/med-data-tools/general-classifier*

## Author contributions

Conceptualization – FD, JZ, NC, JH

Methodology, technical implementation – FD, JZ, RG

Methodology, creation of datasets – PW, MW, SA, JH

Statistical Analysis – FD

Writing, original draft preparation – FD, PW, JZ, PMP, NC, MW, MSc, MSh

Writing, illustrations – FD, PW, IF

Writing, review and editing – All authors

Project administration – FD, NC, JH

## Acknowledgements

Not applicable.

