## Supplementary Table 1 for "Application of a general LLM-based classification system to retrieve information about oncological trials"

Supplementary Table 1. Portion of valid responses for the classification task given by the three LLMs run via cloud computing using output analysis via regular expressions instead of constrained generation.

|  | **Mixtral-8x7B-Instruct-v0.1** | **Llama3.1-70B-Instruct** | **Qwen2.5-72B-Instruct** |
| --- | --- | --- | --- |
| Overall | 99.70% | 100.00% | 99.88% |
| Is this a randomized controlled trial (RCT)? | 100.00% | 100.00% | 99.67% |
| Is this an oncological trial? | 100.00% | 100.00% | 100.00% |
| Were patients with localized disease included in this trial? | 97.67% | 100.00% | 99.83% |
| Were patients with distant metastases included in this trial? | 99.83% | 100.00% | 99.83% |
| Is this trial about patients with breast cancer? | 100.00% | 100.00% | 100.00% |
| Is this trial about patients with prostate cancer? | 100.00% | 100.00% | 99.83% |
| Is this trial about patients with anal cancer? | 100.00% | 100.00% | 100.00% |
| Is this trial about patients with metastatic castration resistant prostate cancer (mCRPC)? | 100.00% | 100.00% | 100.00% |
| Does this trial investigate a therapeutic intervention (e.g., drug, biological, others)? | 100.00% | 100.00% | 100.00% |
