## Supplementary Table 2 for "Application of a general LLM-based classification system to retrieve information about oncological trials"

Supplementary Table 2. Classification metrics of different LLMs on the nine questions.

|  | | ***Local Hardware*** | ***Cloud computing*** | | |
| --- | --- | --- | --- | --- | --- |
|  | | **Mixtral-8x7B-Instruct-v0.1** | **Mixtral-8x7B-Instruct-v0.1** | **Llama3.1-70B-Instruct** | **Qwen2.5-72B-Instruct** |
| ***Is this a randomized controlled trial (RCT)?*** | | | | | |
|  | Accuracy | 93.33% | 93.66% | 96.00% | 93.97% |
|  | Precision | 88.98% | 90.99% | 92.12% | 88.58% |
|  | Recall | 97.86% | 95.96% | 100.00% | 100.00% |
|  | F1 | 93.21% | 93.41% | 95.90% | 93.95% |
| ***Is this an oncological trial?*** | | | | | |
|  | Accuracy | 97.00% | 97.89% | 98.55% | 98.67% |
|  | Precision | 99.54% | 98.27% | 96.36% | 97.92% |
|  | Recall | 89.26% | 93.80% | 98.35% | 97.11% |
|  | F1 | 94.12% | 95.98% | 97.34% | 97.51% |
| ***Were patients with localized cancer disease included in this trial?*** | | | | | |
|  | Accuracy | 86.33% | 85.49% | 90.50% | 77.13% |
|  | Precision | 86.85% | 91.67% | 90.38% | 91.62% |
|  | Recall | 95.63% | 88.51% | 97.24% | 75.40% |
|  | F1 | 91.03% | 90.06% | 93.69% | 82.72% |
| ***Were patients with distant metastases included in this trial?*** | | | | | |
|  | Accuracy | 95.17% | 96.67% | 96.50% | 91.49% |
|  | Precision | 98.52% | 99.41% | 99.41% | 99.35% |
|  | Recall | 93.28% | 94.96% | 94.68% | 86.24% |
|  | F1 | 95.83% | 97.13% | 96.99% | 92.33% |
| ***Is this trial about patients with breast cancer?*** | | | | | |
|  | Accuracy | 98.00% | 98.67% | 97.83% | 98.50% |
|  | Precision | 93.79% | 98.52% | 97.01% | 99.24% |
|  | Recall | 97.84% | 95.68% | 93.53% | 94.24% |
|  | F1 | 95.77% | 97.08% | 95.24% | 96.68% |
| ***Is this trial about patients with prostate cancer?*** | | | | | |
|  | Accuracy | 99.33% | 99.50% | 99.33% | 99.33% |
|  | Precision | 95.16% | 96.72% | 98.28% | 96.67% |
|  | Recall | 98.33% | 98.33% | 95.00% | 96.67% |
|  | F1 | 96.72% | 97.52% | 96.61% | 96.67% |
| ***Is this trial about patients with anal cancer?*** | | | | | |
|  | Accuracy | 99.83% | 99.83% | 99.83% | 99.83% |
|  | Precision | 100.00% | 100.00% | 100.00% | 100.00% |
|  | Recall | 66.67% | 66.67% | 66.67% | 66.67% |
|  | F1 | 80.00% | 80.00% | 80.00% | 80.00% |
| ***Is this trial about patients with metastatic castration resistant prostate cancer (mCRPC)?*** | | | | | |
|  | Accuracy | 95.83% | 97.22% | 97.92% | 99.31% |
|  | Precision | 91.80% | 93.44% | 100.00% | 100.00% |
|  | Recall | 98.25% | 100.00% | 94.74% | 98.25% |
|  | F1 | 94.92% | 96.61% | 97.30% | 99.12% |
| ***Does this trial investigate a therapeutic intervention (e.g., drug, biological, others)?*** | | | | | |
|  | Accuracy | 93.75% | 95.31% | 95.31% | 95.31% |
|  | Precision | 93.44% | 95.00% | 95.00% | 95.00% |
|  | Recall | 100.00% | 100.00% | 100.00% | 100.00% |
|  | F1 | 96.61% | 97.44% | 97.44% | 97.44% |
